# No impact of smoking status on breast cancer tumor infiltrating lymphocytes, response to neoadjuvant chemotherapy and prognosis

**DOI:** 10.1101/2020.06.05.20123273

**Authors:** Vanille Simon, Lucie Laot, Enora Laas, Sonia Rozette, Julien Guerin, Thomas Balezeau, Marion Nicolas, Jean-Yves Pierga, Florence Coussy, Marick Laé, Diane de Croze, Beatriz Grandal, Judith Abecassis, Fabien Reyal, Anne-Sophie Hamy

## Abstract

**INTRODUCTION:** Tobacco use is associated with an increase in breast cancer (BC) mortality. Pathologic complete response rate to neoadjuvant chemotherapy (NAC) is influenced by tumor-infiltrating lymphocytes (TILs) levels and is associated with a better long-term survival outcome. Whether tobacco modifies either tumoral microenvironment such as TIL levels, either pCR rates remains unclear. The aim of our study is to evaluate the impact of smoking status on TIL levels, response to NAC and prognosis for BC patients.

**METHODS:** We retrospectively evaluated pre and post NAC stromal and intra tumoral TIL levels and pCR rates on a cohort of T1-T3NxM0 BC patients treated with NAC between 2002 and 2012 at Institut Curie. Smoking status (current, ever, never smokers) was collected in clinical records. We analyzed the association between smoking status, TIL levels, pCR rates and survival outcomes among the whole population, and after stratification by BC subtype.

**RESULTS:** A total of 956 BC patients with available smoking status information were included in our analysis [current smokers, n=179 (18.7%); ever smokers, n=154 (16.1%) and never smokers, n = 623 (65.2%)]. Median pre-NAC TIL levels, pCR rates, or median post-NAC TIL levels were not significantly different according to smoking status, neither in the whole population, nor after stratification by BC subtype. With a median follow-up of 101.4 months, relapse free survival (RFS) and overall survival (OS) were not significantly different by smoking status.

**CONCLUSION:** In this study, we did not find any significant effect of tobacco use on pre and post NAC TILs nor response to NAC and. Though our data seem reassuring, BC treatment should still be considered as a window of opportunity to offer BC patients accurate smoking cessation interventions.

## BACKGROUND

Neoadjuvant chemotherapy (NAC) is currently prescribed for patients with locally advanced breast cancers (BC) (T3-T4), triple negative (TNBC), *HER2*-positive status or positive nodal status. Beyond increasing breast-conserving surgery rates[1], it also serves as an *in vivo* chemosensitivity test and the analysis of residual tumor burden may help understanding resistance to treatment[2]. Moreover, pathological complete response (pCR) after NAC has been associated with a better long term survival outcome [1][3].

Denkert *et al*. first evidenced that the amount of stromal **immune infiltration** was positively associated with pathological complete response (pCR) after NAC[4]. These results were recently confirmed on a pooled analysis of a large cohort of 3771 patients receiving NAC from German Breast Group[5], showing that the relationship between TIL levels and pCR translated into a disease free survival advantage in *HER2*-positive and TNBC tumors. The drivers of immunosurveillance derive from both tumor-intrinsic characteristics, and extrinsic factors related to the host or the environment[6–8]. Among endogenous tumor characteristics, BC subtype and proliferative patterns are the main factors associated with immune infiltration. Extrinsic factors including notably environment (tobacco, alcohol), nutritional factors and diet have been studied less extensively. The identification of factors associated with changes in the microenvironment is of particular interest, as lifestyle factors are actionable and their changes could theoretically improve prognosis.

Tobacco smoking is known to be associated with an increased risk of several cancer types, including larynx, oropharynx, esophagus, lung, bladder, kidney, urinary tract, cervix, colorectum and gastrointestinal tract, and acute myeloid leukemia[9]. Tobacco deregulates many biological pathways, and induces inflammation, impaired immune function and DNA damage[10, 11], leading to an increase of tumor proliferation, invasion and angiogenesis. Regarding BC, tobacco is associated with post-operative complications[18–20], radiation-induced toxicities[21–23], and altered quality of life[24]. A recent meta-analysis[12] on 39,725 patients reported that smoking increased the risk of breast cancer, all-cause and specific mortality, and that BC patients who smoke have a higher risk of second primary cancer[13], and ipsilateral lung cancer [14–17] when combined with radiotherapy. However, there is few data on the relationship between cigarette smoking, immune infiltration and response to neoadjuvant chemotherapy in BC.

The aim of our study is to analyze the relationships between smoking status at BC diagnosis, tumor infiltrating lymphocytes, response to NAC and prognosis on a large cohort of BC patients treated with NAC.

## METHODS

### Patients and tumors

We analyzed a cohort of 1199 T1–3NхM0 patients with invasive breast carcinoma (NEOREP Cohort, CNIL declaration number 1547270) treated at Institut Curie (Paris and Saint Cloud, France), between 2002 and 2012. We included patients with unilateral, non-recurrent, noninflammatory, non-metastatic tumors for whom NAC was indicated. Every patient received NAC, followed by surgery, radiotherapy and endocrine therapy when indicated. The study was approved by the Breast Cancer Study Group of Institut Curie and was conducted according to institutional and ethical rules regarding research on tissue specimens and patients. Written informed consent from the patients was not required by French regulations.

### Tobacco smoking

Data regarding history of smoking was collected retrospectively in May 2018 in clinical records (either in oncology or gynecology first consultation, either in anesthesiology consultations) for the purpose of the current study. We defined ***current smokers*** as active smokers at the time of BC diagnosis, ***ever smokers*** as patients with a prior history of smoking having stopped before BC diagnosis, and ***never smokers*** as patients who had never smoked. We also documented smoking intensity through the use of pack-years, which is a measurement unit calculated by multiplying the number of packs of cigarettes smoked per day by the number of years the person has smoked. Heavy smokers were defined as 20 or more pack-years.

### Treatments

Patients were treated according to national guidelines. NAC regimens changed over time (anthracycline-based regimen or sequential anthracycline-taxane regimen), and trastuzumab was used in an adjuvant and/or neoadjuvant setting since 2005 for *HER2*-positive breast cancer. Surgery was performed four to six weeks after the end of chemotherapy. Most patients(98.2%, n=1127) received adjuvant radiotherapy. Endocrine therapy (tamoxifen, aromatase inhibitor, and/or GnRH agonists) was prescribed when indicated.

### Tumor samples

BC tumors were classified into subtypes (TNBC, *HER2*-positive, and luminal) on the basis of immunohistochemistry. ER and PR status were determined as follows. Tissue sections were rehydrated, and antigen retrieval was carried out in citrate buffer (10 mM, pH 6.1). The sections were then incubated with antibodies against ER (clone 6F11, Novocastra, Leica Biosystems, Newcastle, UK; 1/200) and PR (clone 1A6, Novocastra, 1/200). Antibody binding was detected with Vectastain Elite ABC peroxidase-conjugated mouse IgG kit (Vector, Burlingame, CA, USA), with diaminobenzidine (Dako A/S, Glostrup, Denmark) as chromogen. Positive and negative controls were included in each run. According to French recommendations, cases were considered positive for ER and PR if at least 10% of tumor nuclei were stained[25]. Tumors were considered hormone receptor (HR)-positive when positive for either ER or PR (referred to hereafter as “luminal”), and HR-negative when negative for both ER and PR. *HER2* expression was determined by immunohistochemistry using a monoclonal anti-*HER2* antibody (CB11, Novocastra, New-Castle, UK; 1/800). Scoring was performed according to American Society of Clinical Oncology (ASCO)/College of American Pathologists (CAP) guidelines[26]. Scores 3+ were reported as positive, scores 0/1+ as negative. Tumors with scores 2+ were tested by fluorescence *in situ* hybridization (FISH). FISH was performed using a *HER2*-gene-specific probe and a centromeric probe for chromosome 17 (PathVysion HER-2 DNA Probe kit, Vysis-Abbott, Abbott Park, IL, USA) according to manufacturers’ instructions. *HER2* gene amplification was defined according to ASCO/CAP guidelines[26]. An average of 40 tumor cells per sample was evaluated and mean *HER2* signals per nuclei were calculated. A *HER2*/CEN17 ratio ≥ 2 was considered positive, and a ratio < 2 was considered negative[26].

### Pathological review

Pretreatment core needle biopsy specimens and the corresponding post-NAC surgical specimens were reviewed independently by two experts in breast diseases. Formalin-fixed paraffin-embedded (FFPE) tumor tissue samples were studied. Pre and post-NAC stromal (str) and intra-tumoral (IT) TILs were reviewed between January 2015 and March 2017 according to the recommendations of the international TILs Working Group [27, 28]. Further details on TILs review are available in [29].

Response to treatment was retrospectively reviewed and was assessed by: (i) pathological complete response, defined as the absence of residual invasive cancer in the breast and axillary nodes (ypT0/ ypN0) after neoadjuvant chemotherapy [30]; (ii) residual cancer burden (RCB) indices as described by Symmans [31], with the web-based calculator freely available via the Internet (https://www.mdanderson.org/breastcancer_RCB). pCR corresponded to an RCB 0 index. Further details on RCB review are available in ref [32].

### Study endpoints

The aims of the study were to analyze the association between smoking status at diagnosis and: (i) Pre and post NAC str TILs; (ii) response to NAC assessed by pathological complete response (pCR) and RCB index; (iii) prognosis assessed by relapse-free survival (RFS). Relapse-free survival (RFS) was defined as the time from surgery to death, loco-regional recurrence or distant recurrence, whichever occurred first. Overall survival (OS) was defined as the time from surgery to death. For patients for whom none of these events were recorded, we censored data at the time of last known contact. Survival cutoff date analysis was February 1^st^, 2019.

### Statistical analysis

The population was described in terms of frequencies for qualitative variables or medians and associated range for quantitative variables. Pre- and post-NAC TIL levels, and RCB index were analyzed as continuous variables. All analyses were performed on the whole population and after stratification by BC subtype. TIL levels and qualitative variables in classes were compared with ANOVA or Mann Whitney tests, when appropriate. Comparisons of proportion of samples were investigated by chi-squared and Fisher tests.

Factors predictive of pCR were introduced into a univariate logistic regression model. A multivariate logistic model was then implemented. Covariates selected for multivariate analysis were those with a p-value likelihood ratio test below 0.05 in univariate analysis. Survival was described using Kaplan-Meier estimate and comparison between survival curves was performed with the Log-rank test. Estimation of hazard ratios (HR) and their associated 95% confidence interval (CI) was carried out using the Cox proportional hazard model. Significance threshold was 5%. Analyses were performed with R software, version 3.1.2.

## RESULTS

### Baseline patients’ and tumors’ characteristics

Among 1199 women included in the cohort, data regarding smoking status were missing for 243 patients (19%), leaving 956 patients for the current analyses. In the whole population, mean age was 48 years-old at diagnosis, and mean body mass index (BMI) was 24 kg/m^2^ (range 16–47). Most of BCs were in T2 stage (66.2%), grade III (61%), and 550 patients(57.6%) had positive axillary nodes. Patient’s repartition by subtype was as follows: luminal (n=410, 42.9%), TNBC (n=305, 31.9%), *HER2*-positive (n=241; 25.2%) (Table 1).

**Table 1:**
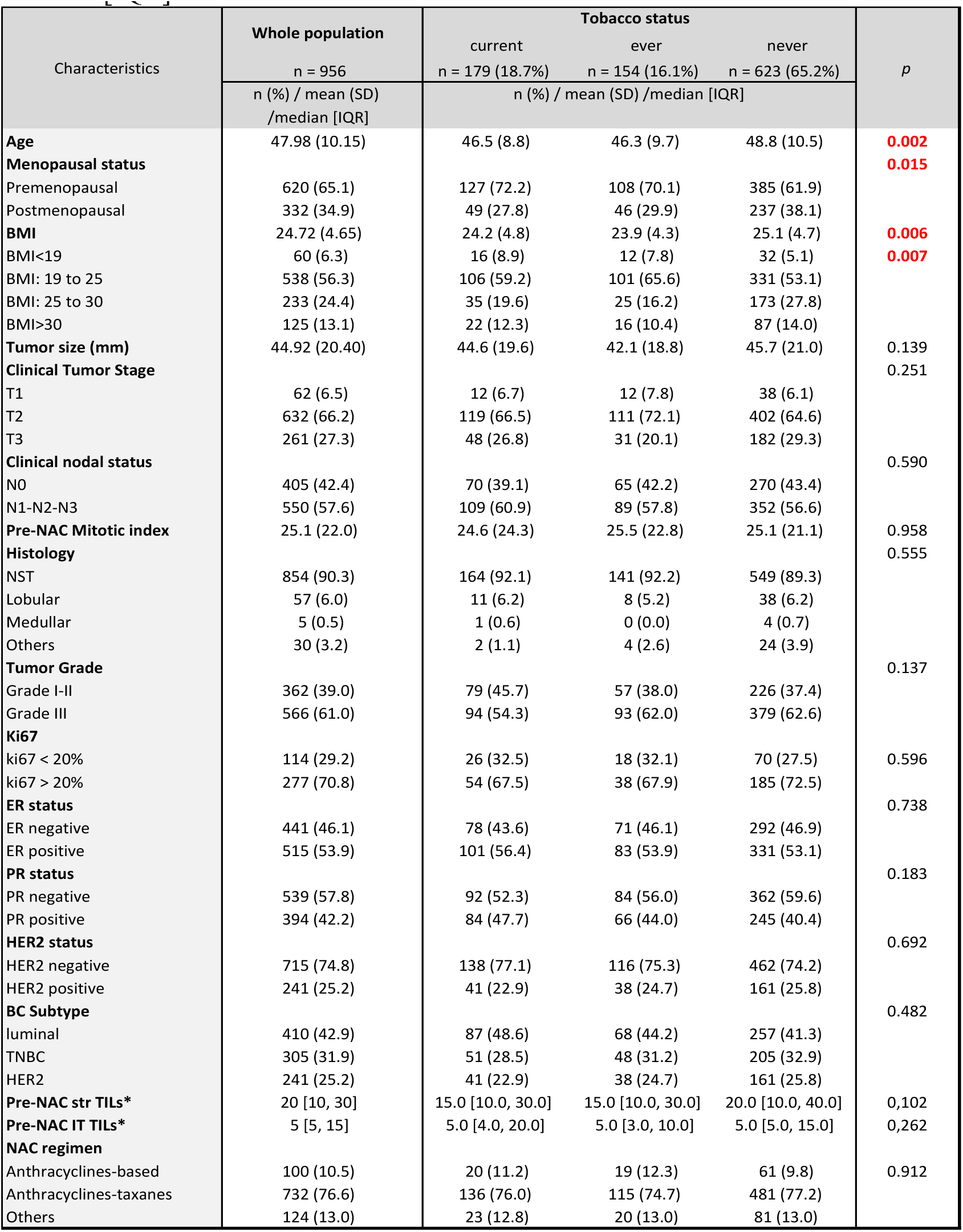
Patients’ characteristics among the whole population and according to tobacco status. Missing data: Menopausal status, n=4; Tumor size (mm), n=1; Clinical tumor stage, n=1; Clinical nodal status, n=1; Mitotic index, n=338; Tumor Grade, n=28; ki67, n=565; Histology, n=10; PR status, n=23; Pre-NAC str TILs, n=324; Pre-NAC IT TILs, n=324 Abbreviations: BMI=body mass index; NST = no special type; ER = estrogen receptor; PR=progesterone receptor; TNBC=Triple Negative Breast Cancer; NAC=neoadjuvant chemotherapy; str=stromal; IT= intratumoral. The “n” denotes the number of patients. In case of categorical variables, percentages are expressed between brackets. In case of continuous variables, mean value is reported, with standard deviation (SD) between brackets. In case of nonnormal continuous variables*, median value is reported, with interquartile range between brackets [IQR].

| Characteristics | Whole population | Tobacco status |  |  | p |
| --- | --- | --- | --- | --- | --- |
|  | n = 956 | current<br>n = 179 (18.7%) | ever<br>n = 154 (16.1%) | never<br>n = 623 (65.2%) |  |
|  | n (%) / mean (SD)<br>/median [IQR] | n (%) / mean (SD) /median [IQR] |  |  |  |
| Age | 47.98 (10.15) | 46.5 (8.8) | 46.3 (9.7) | 48.8 (10.5) | 0.002 |
| Menopausal status |  |  |  |  | 0.015 |
| Premenopausal | 620 (65.1) | 127 (72.2) | 108 (70.1) | 385 (61.9) |  |
| Postmenopausal | 332 (34.9) | 49 (27.8) | 46 (29.9) | 237 (38.1) |  |
| BMI | 24.72 (4.65) | 24.2 (4.8) | 23.9 (4.3) | 25.1 (4.7) | 0.006 |
| BMI<19 | 60 (6.3) | 16 (8.9) | 12 (7.8) | 32 (5.1) | 0.007 |
| BMI: 19 to 25 | 538 (56.3) | 106 (59.2) | 101 (65.6) | 331 (53.1) |  |
| BMI: 25 to 30 | 233 (24.4) | 35 (19.6) | 25 (16.2) | 173 (27.8) |  |
| BMI>30 | 125 (13.1) | 22 (12.3) | 16 (10.4) | 87 (14.0) |  |
| Tumor size (mm) | 44.92 (20.40) | 44.6 (19.6) | 42.1 (18.8) | 45.7 (21.0) | 0.139 |
| Clinical Tumor Stage |  |  |  |  | 0.251 |
| T1 | 62 (6.5) | 12 (6.7) | 12 (7.8) | 38 (6.1) |  |
| T2 | 632 (66.2) | 119 (66.5) | 111 (72.1) | 402 (64.6) |  |
| T3 | 261 (27.3) | 48 (26.8) | 31 (20.1) | 182 (29.3) |  |
| Clinical nodal status |  |  |  |  | 0.590 |
| N0 | 405 (42.4) | 70 (39.1) | 65 (42.2) | 270 (43.4) |  |
| N1-N2-N3 | 550 (57.6) | 109 (60.9) | 89 (57.8) | 352 (56.6) |  |
| Pre-NAC Mitotic index | 25.1 (22.0) | 24.6 (24.3) | 25.5 (22.8) | 25.1 (21.1) | 0.958 |
| Histology |  |  |  |  | 0.555 |
| NST | 854 (90.3) | 164 (92.1) | 141 (92.2) | 549 (89.3) |  |
| Lobular | 57 (6.0) | 11 (6.2) | 8 (5.2) | 38 (6.2) |  |
| Medullar | 5 (0.5) | 1 (0.6) | 0 (0.0) | 4 (0.7) |  |
| Others | 30 (3.2) | 2 (1.1) | 4 (2.6) | 24 (3.9) |  |
| Tumor Grade |  |  |  |  | 0.137 |
| Grade I-II | 362 (39.0) | 79 (45.7) | 57 (38.0) | 226 (37.4) |  |
| Grade III | 566 (61.0) | 94 (54.3) | 93 (62.0) | 379 (62.6) |  |
| Ki67 |  |  |  |  |  |
| ki67 < 20% | 114 (29.2) | 26 (32.5) | 18 (32.1) | 70 (27.5) | 0.596 |
| ki67 > 20% | 277 (70.8) | 54 (67.5) | 38 (67.9) | 185 (72.5) |  |
| ER status |  |  |  |  | 0.738 |
| ER negative | 441 (46.1) | 78 (43.6) | 71 (46.1) | 292 (46.9) |  |
| ER positive | 515 (53.9) | 101 (56.4) | 83 (53.9) | 331 (53.1) |  |
| PR status |  |  |  |  | 0.183 |
| PR negative | 539 (57.8) | 92 (52.3) | 84 (56.0) | 362 (59.6) |  |
| PR positive | 394 (42.2) | 84 (47.7) | 66 (44.0) | 245 (40.4) |  |
| HER2 status |  |  |  |  | 0.692 |
| HER2 negative | 715 (74.8) | 138 (77.1) | 116 (75.3) | 462 (74.2) |  |
| HER2 positive | 241 (25.2) | 41 (22.9) | 38 (24.7) | 161 (25.8) |  |
| BC Subtype |  |  |  |  | 0.482 |
| luminal | 410 (42.9) | 87 (48.6) | 68 (44.2) | 257 (41.3) |  |
| TNBC | 305 (31.9) | 51 (28.5) | 48 (31.2) | 205 (32.9) |  |
| HER2 | 241 (25.2) | 41 (22.9) | 38 (24.7) | 161 (25.8) |  |
| Pre-NAC str TILs* | 20 [10, 30] | 15.0 [10.0, 30.0] | 15.0 [10.0, 30.0] | 20.0 [10.0, 40.0] | 0,102 |
| Pre-NAC IT TILs* | 5 [5, 15] | 5.0 [4.0, 20.0] | 5.0 [3.0, 10.0] | 5.0 [5.0, 15.0] | 0,262 |
| NAC regimen |  |  |  |  |  |
| Anthracyclines-based | 100 (10.5) | 20 (11.2) | 19 (12.3) | 61 (9.8) | 0.912 |
| Anthracyclines-taxanes | 732 (76.6) | 136 (76.0) | 115 (74.7) | 481 (77.2) |  |
| Others | 124 (13.0) | 23 (12.8) | 20 (13.0) | 81 (13.0) |  |

At BC diagnosis, 179 patients were current smokers (18.7%), 154 were ever smokers (16.1%) and 623 women were never smokers (65.2%). Among 333 patients with a previous tobacco history (34.8%), 23% (n=77) were heavy smokers > 20 pack-years. The median pack-year was 15 (range 1–100). Patients’ characteristics according to smoking status are described in Table 1. Current and ever smokers were statistically younger (46.4 *versus* 48.8 y.o., *p* = 0.002) and thinner (BMI 24 *versus* 25, *p* = 0.006) than never smokers. Conversely, no tumor characteristic was statistically different according to smoking status, among tumor size, histology, mitotic index, ER, PR, *HER2* or nodal status (Table 1).

Pre-NAC str TIL levels were available for 632 patients and were not significantly different between current, ever, and never smokers (15%, 15% and 20% respectively, *p =* 0.1, Figure 1). This was true in the whole population (Figure 1A) and after stratification by BC subtype (Figure 1B). Similarly, IT TIL levels were not significantly different according to smoking status in the whole population (Figure 1C current: 5%, ever: 5% and never 5%, *p =* 0.26), neither were they after stratification by BC subtype (Figure1D). Among current smokers, no difference was found in stromal or IT TIL levels according to smoking intensity (Supplemental Figure 1).

**Figure 1:**
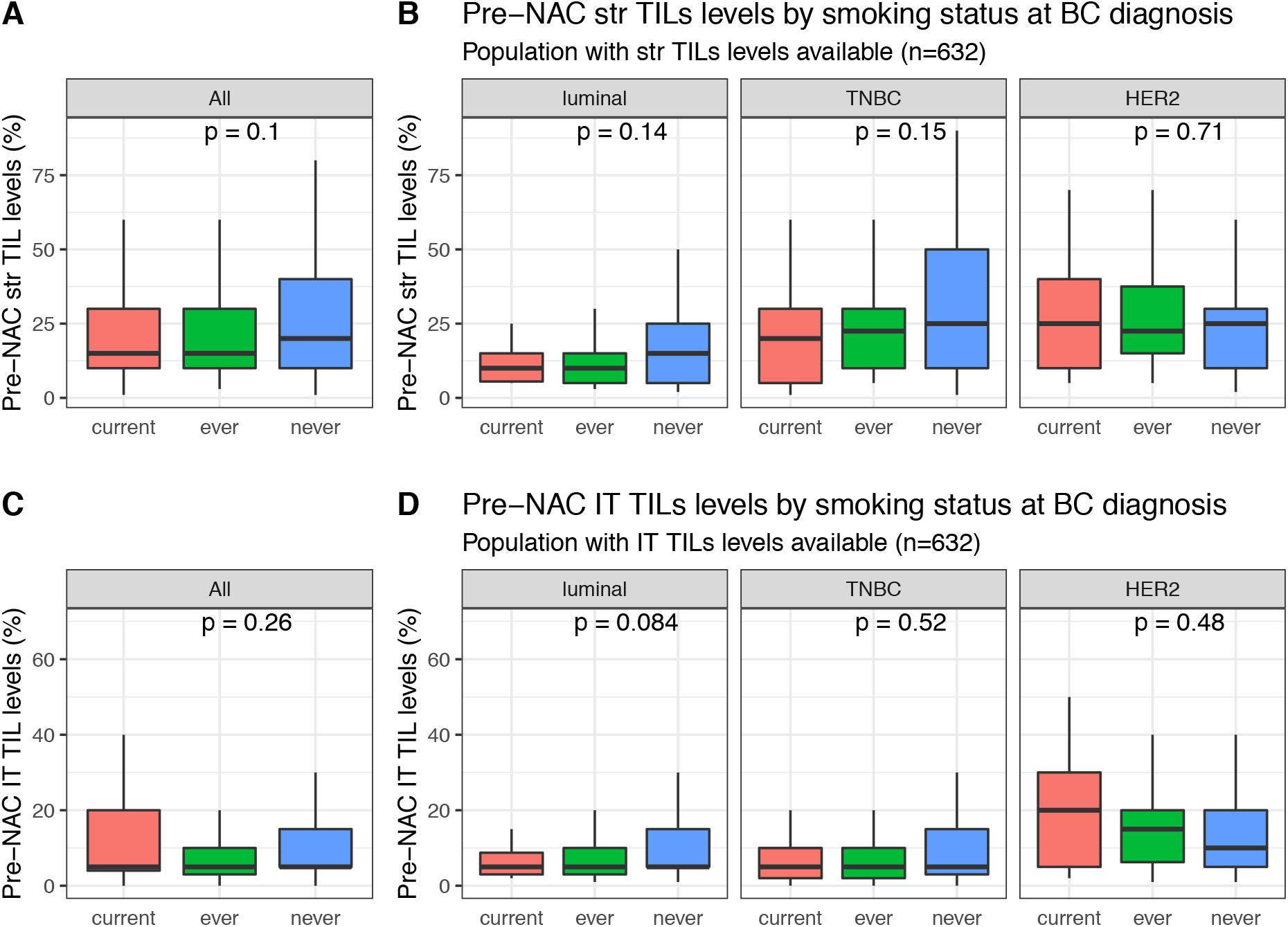
Pre-NAC TIL levels at BC diagnosis according to smoking status: Pre-NAC str TIL levels in the whole population (A), Pre-NAC str TIL levels by BC subtype (B), Pre-NAC IT TIL levels in the whole population (C), Pre-NAC IT TIL levels by BC subtype (D).

### Response to treatment and post-NAC immune infiltration

After NAC, the pathological complete response rate was 25.8% (247/956) and this rate was different by BC subtype (luminal: 6.8% (28/410); TNBC: 39.7% (121/305), *HER2*-positive:40.7% (98/241), p<0.001). pCR rates were not significantly different in current, ever or never smokers (20.7%, 29.2% and 26.5% respectively, *p =* 0.17, Table 2, Figure 2A). Similar results were found after stratification by pathological subtype (luminal: *p =* 0.63; TNBC: *p =* 0.27; *HER2*-positive: *p =* 0.22, figure 1B). In the same vein, no difference was seen regarding RCB index (Supplemental Figure 2).

**Table 2:**
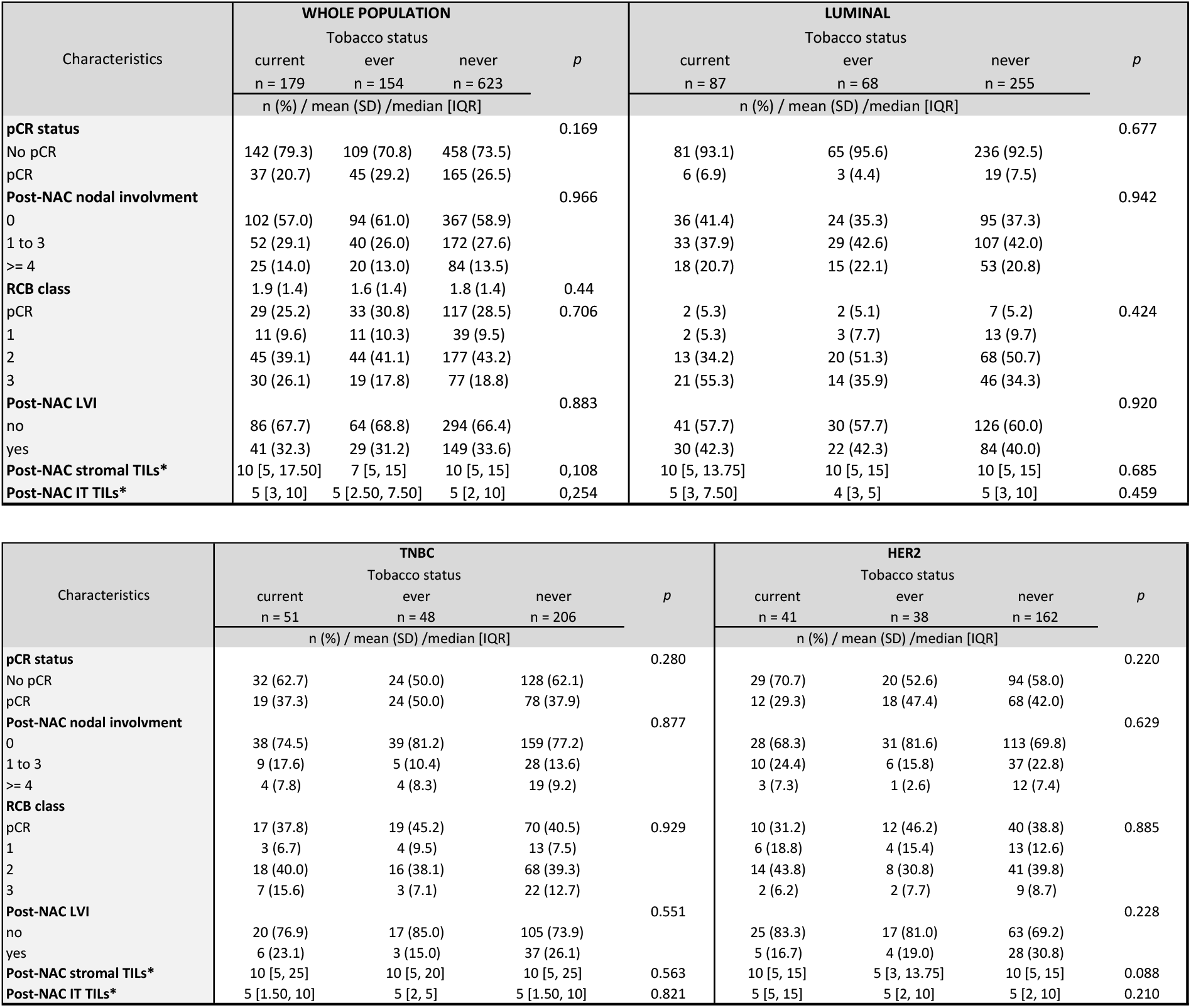
Post-NAC TILs, pCR and RCB class according to smoking status. Missing data: RCB class, n= 80; Post-NAC LVI, n=99; Post-NAC str TILs, n=80; Post-NAC IT TILS, n=145. Abbreviations: pCR = pathological Complete Response; NAC = Neoadjuvant Chemotherapy; RCB = Residual Cancer Burden; LVI = Lympho-Vascular Involvement; str = stromal; IT = intratumoral.The “n” denotes the number of patients. In case of categorical variables, percentages are expressed between brackets. In case of continuous variables, mean value is reported, with standard deviation (SD) between brackets. In case of nonnormal continuous variables*, median value is reported, with interquartile range between brackets [IQR].

**Figure 2:**
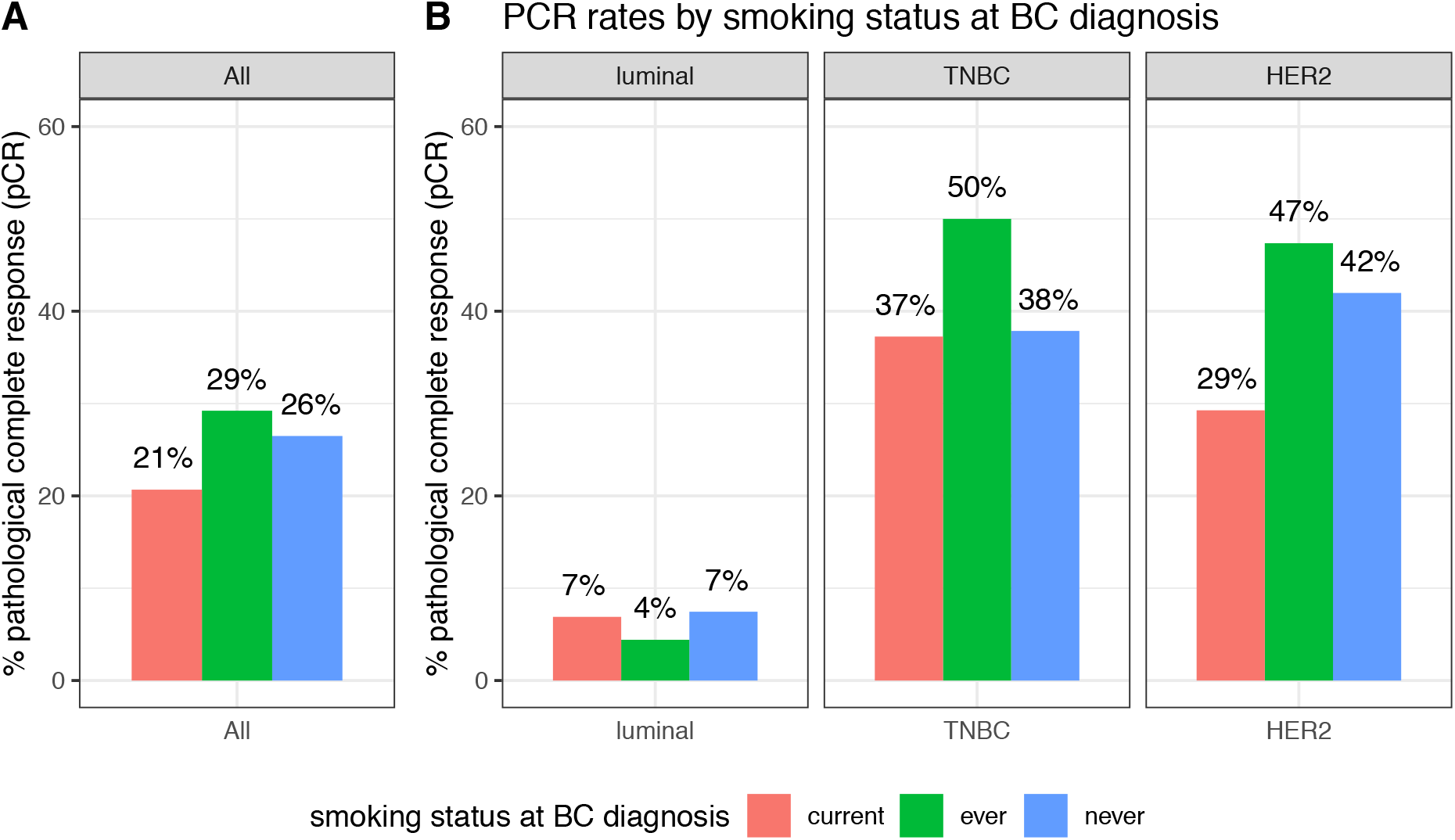
pCR rates according to smoking status at BC diagnosis, in the whole population (A) and by BC subtype (B).

Post-NAC str and IT TILs were available for 632 and 429 patients, respectively. Post-NAC median str TIL levels were not significantly different between current, ever and never smokers in the whole population (10%, 7% and 10% respectively, p = 0.108, Figure 3A) nor in each BC subtype (Figure 3C). Similar results were found for post-NAC IT TIL levels both in the whole population (5%, 5% and 5% respectively, p = 0.254) and after stratification by BC subtype (Figure 3C and 3D respectively).

**Figure 3:**
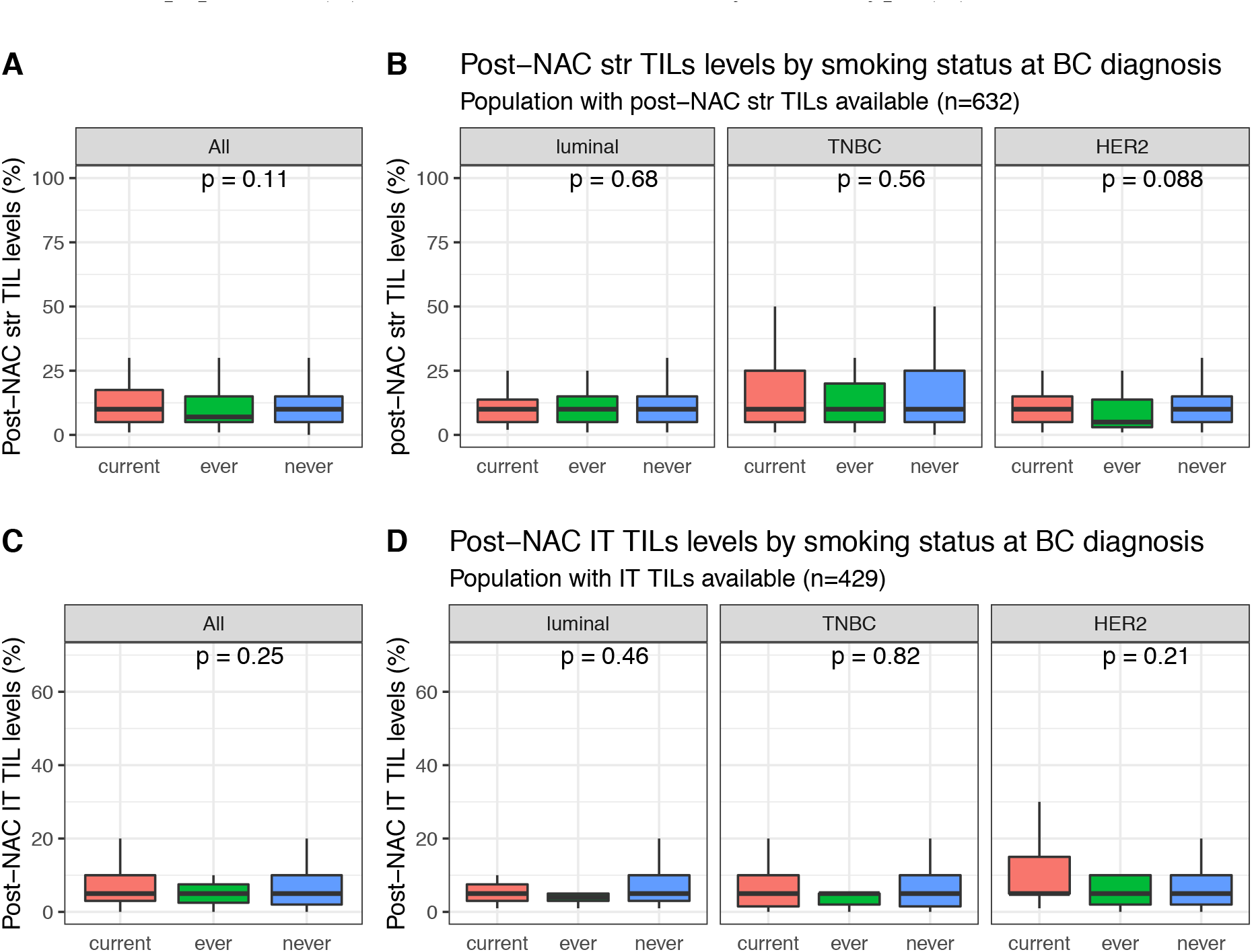
Post NAC TIL levels according to smoking status: Post-NAC str TIL levels in the whole population (A), Post-NAC str TIL levels by BC subtype (B), Post-NAC IT TIL levels in the whole population (C), Post-NAC IT TIL levels by BC subtype (D).

### Survival outcomes

With a median follow-up of 101.4 months, 293 patients experienced relapse, and 173 died. Tobacco smoking was not significantly associated with RFS, neither in the whole population, nor after stratification by BC subtype (Figure 4). Similar results were observed for overall survival, where tobacco smoking at diagnosis had no impact at all on mortality in the whole population (Figure 5). After stratification by BC subtype, patients with luminal BC who were current smokers had a worse overall survival when compared with ever or never smokers (*P_interaction_* smoking status /BC subtype = 0.11), but this association was no longer significant after multivariate analysis (Supplemental Table 1).

**Figure 4:**
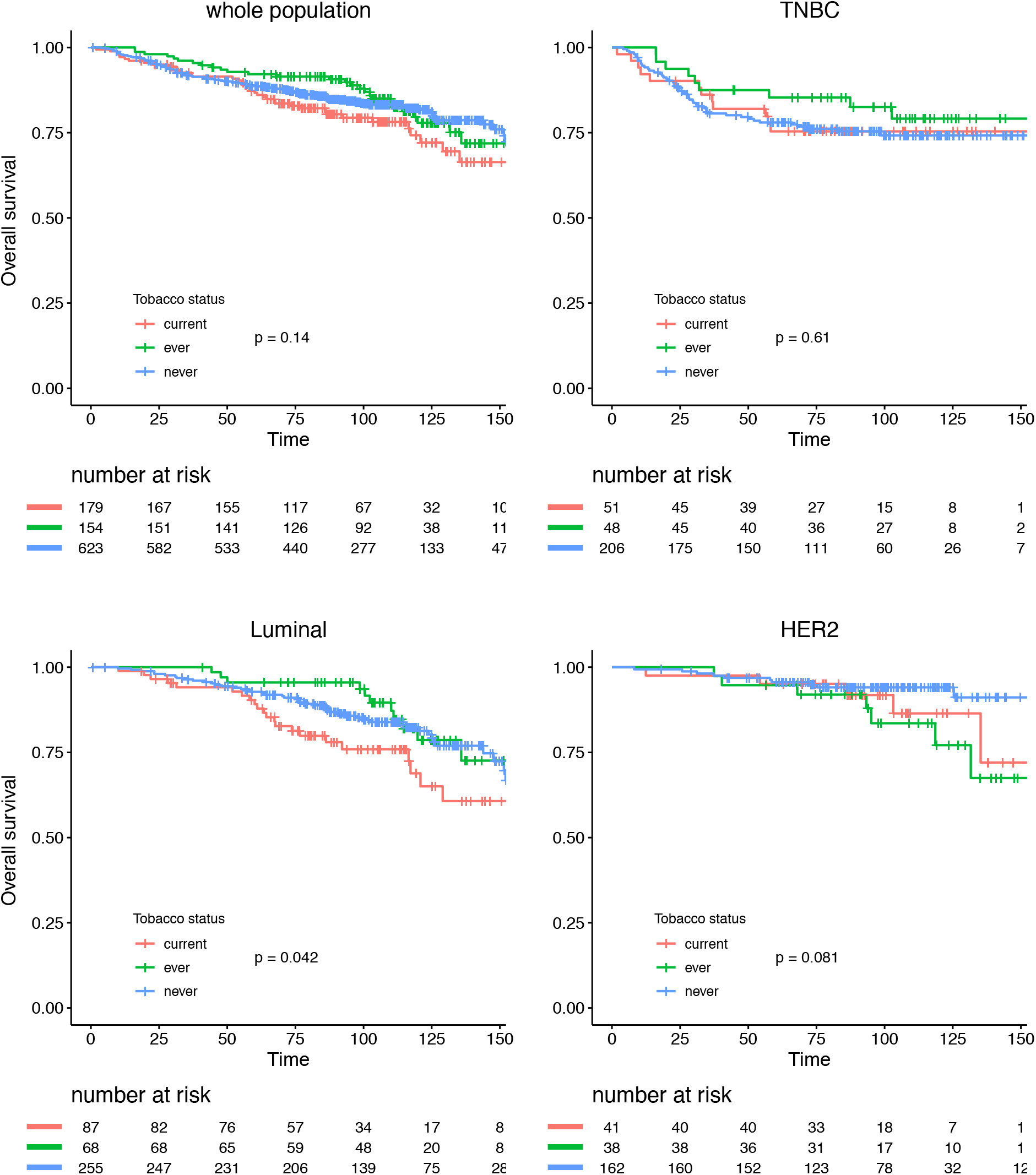
Relapse-free survival according to smoking status in the whole population and by BC subtype.

**Figure 5:**
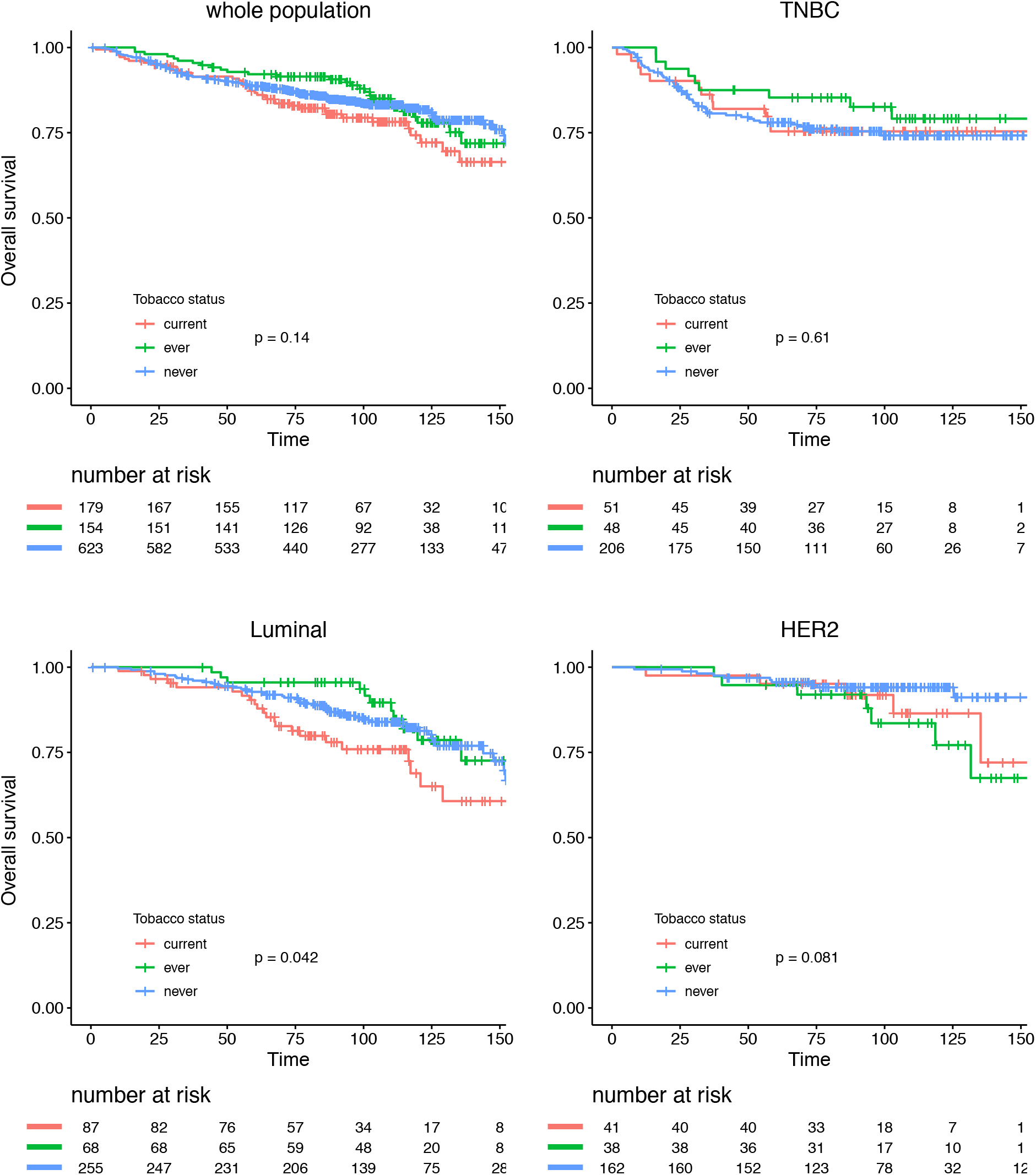
Overall survival according to smoking status in the whole population and by BC subtype.

## Discussion

In this retrospective study evaluating the association between smoking status and oncologic outcomes, we found no significant association between smoking status, pre- and post-NAC immune infiltration, response to treatment or prognosis in a large cohort of BC patients treated with NAC. These findings are of interest, because a significant increase in female tobacco consumption has been observed worldwide over the last century[33], and whether tobacco use could affect response to chemotherapy and prognosis has been barely explored so far.

To our knowledge, only one previous study evaluated the relationship between smoking and the immune microenvironment in BC. On a retrospective study assessing the relationship between tobacco use, TIL levels and pCR in 149 women with *HER2-*positive and TNBCs, Takada and colleagues [34] found that TIL levels and pCR rates were significantly higher in the high-smokers group (defined by more than 2.5 pack-year) than in the low smokers group (TIL levels: 72.1% *versus* 56.6%, *p=* 0.043; pCR 62.8% *versus* 44.3%, p=0.042). With a larger study sample, our results do not support these findings.

Conversely, tobacco use was reported to have a negative impact on survival in patients treated with endocrine therapy. In a cohort of 1116 patients, Persson *et al*. [35] showed an increased risk of recurrences (HR 2.97, 95%CI [1.81–9.72]), distant metastasis (HR 4.19; 95%CI [1.81– 9.72]) and death (HR 3.52; 95%CI [1.59–7.81]) among aromatase-inhibitors treated patients who smoked compared with non-smokers. However, there was no effect in patients treated with Tamoxifene.

In organs directly affected by tobacco, several studies [36–40] showed that perpetuation of tobacco use during radiotherapy was associated with a reduction of its effectiveness and overall survival. In head and neck cancers, Chen *et al*. [36] reported that active smokers during radiotherapy had a significantly inferior 5-year overall survival (23% *vs*. 55%, *p*<0.05), locoregional control (58% *vs*. 69%, *p*<0.005), and disease-free survival (42% vs. 65%, *p*<0.05) when compared with the former smokers who had quit before radiation therapy. In lung cancers, Videtic *et al*. [40] showed a significantly better 5-year overall survival in weaned patients, compared to active smokers during radiotherapy (8.9% *versus* 4% respectively, *p*=0.0017).

A strong biological and preclinical rationale could have supported the working hypothesis that immune response to tumor and sensitivity to chemotherapy may be impaired - or modified - by tobacco. Indeed, nicotine deregulates cell proliferation, apoptosis, migration, invasion, angiogenesis, inflammation, epithelial-mesenchymal transition and cell-mediated immunity in a wide variety of cells, leading to enhanced tumor growth and metastasis [10, 41, 42]. The effects of nicotine are usually mediated through the nicotinic acetylcholine receptors (nAChRs) [43–45], that in turn activate several oncogenic pathways as Ras/Raf/MAPK and PI3K/AKT cascades. Smoking has been reported to induce chemoresistance *in vitro*, in colorectal [46], bladder [36], pancreatic [47] and nasal [48] cancers. Nicotine suppressed chemotherapeutic-induced apoptosis of breast cancer cells *in vitro* [49], via the signaling cascade involving STAT3, galectine-3, and a nicotinic acetylcholine receptor. Tobacco use may also modify pharmacodynamics of anticancer agents [50–52]. In lung cancer, smokers receiving erlotinib or camptothecine showed a rapid clairance requiring a higher dose to reach equivalent systemic effect than never smokers [53].

Recent analyses of cancer genomes have highlighted an association between mutational processes and the catalogue of somatic single nucleotide substitutions observed in a tumor sample. In particular specific processes are characterized by preferential substitution types, and sequence context (defined by the two flanking nucleotides) https://www.biorxiv.org/content/biorxiv/early/2018/05/15/322859.full.pdf

In a systematic analysis of 5243 cancers of types for which tobacco smoking confers an elevated risk (breast cancer was not included), signature 4 - the main marker of tobacco-smoking-caused mutations was only identified in cancers from tissues directly exposed to tobacco smoke - suggesting that increased risk associated with tobacco may be mediated by mechanisms other than an increased mutation load as previously believed https://science.sciencemag.org/content/354/6312/618; immunity could be a good candidate, that could be further explored in other cancer types without direct exposition to smoke, such as pancreatic or bladder.

Finally, tobacco may also play a role in inflammation and deregulation of innate and adaptative immunity [54, 55] and notably affects T-cell lymphocytes functions [54, 56]. In non-small-cell lung cancers, Deng et al. showed that current/ever smokers had higher PDCD1 and CTLA-4 expression in tumor tissues, compared with never smokers (*PDCD1* median 142 vs. 36, *p* < 0.01; *CTLA-4* median 152 vs. 59, *p* < 0.01)[57]. In non-small-cell lung cancer, both Nivolumab[58] and MPDL3280A[59] (a PD-L1 antibody) have been reported to be more active in current/ever smokers than in never-smokers.

However, despite such abundant scientific rationale, our study provides reassuring data on the impact of smoking on BC outcomes in the neoadjuvant setting. Strengths of this study include the large sample size, the upfront evaluation of both pre- and post-NAC IT and stromal TILs, and the long-term follow-up. Limitations should also be considered, such as the absence of collection of smoking habits after diagnosis.

Finally, in front of the well-known benefits of weaning on healing, quality of life, and overall survival, breast cancer treatment and follow-up should be considered as windows of opportunity to address tobacco use and to offer patients accurate smoking cessation alternatives.

## Data Availability

The authors confirm that the data supporting the findings of this study are available within the article [and/or] its supplementary material.

---

**Supplementary Table 1:**
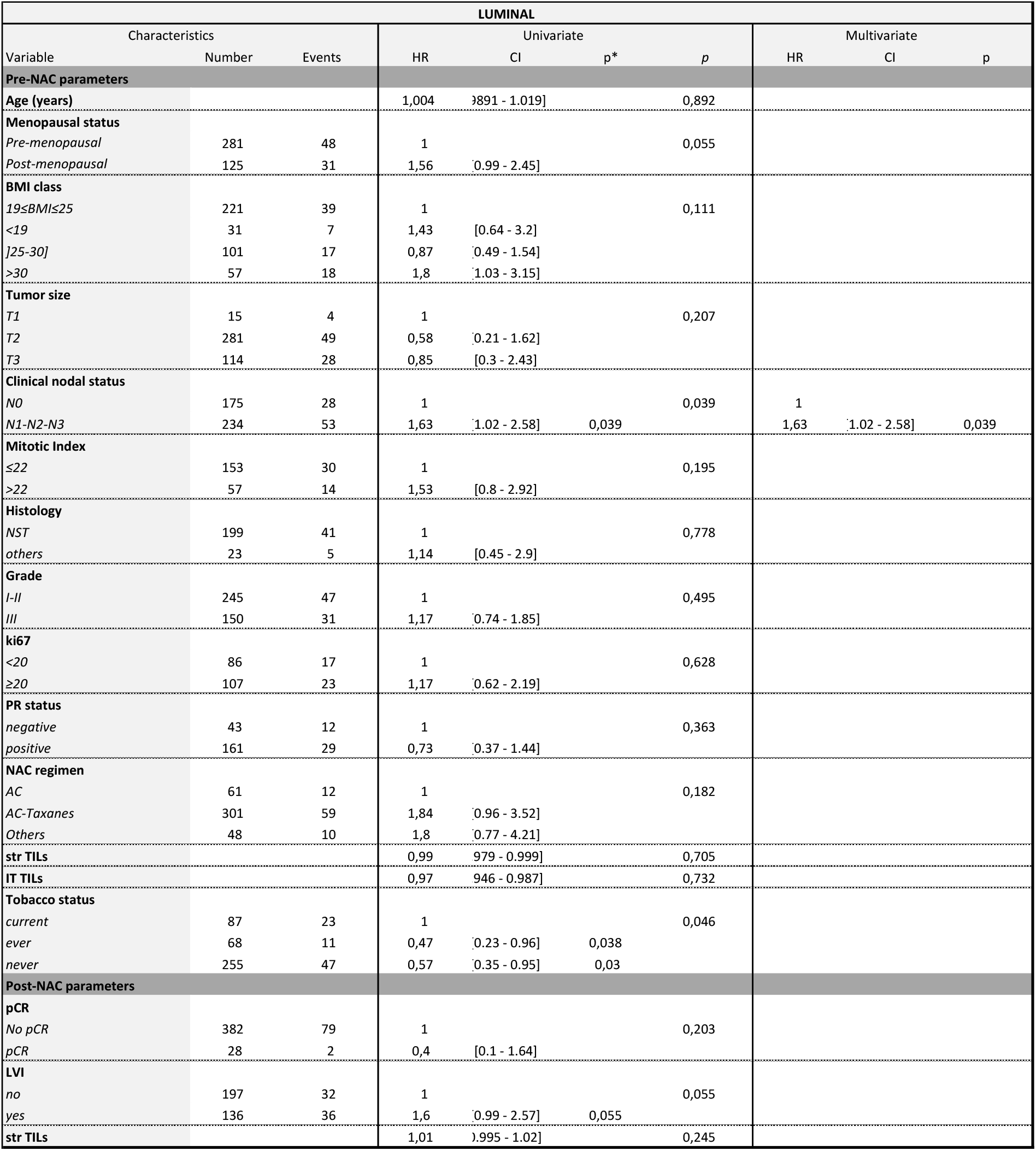
Univariate and multivariate analysis on overall survival in the luminal BC population. Abbreviations: pCR = pathological Complete Response; NAC = Neoadjuvant Chemotherapy; RCB = Residual Cancer Burden; LVI = Lympho-Vascular Involvement; str = stromal; IT = intratumoral

**Supplementary Figure 1:**
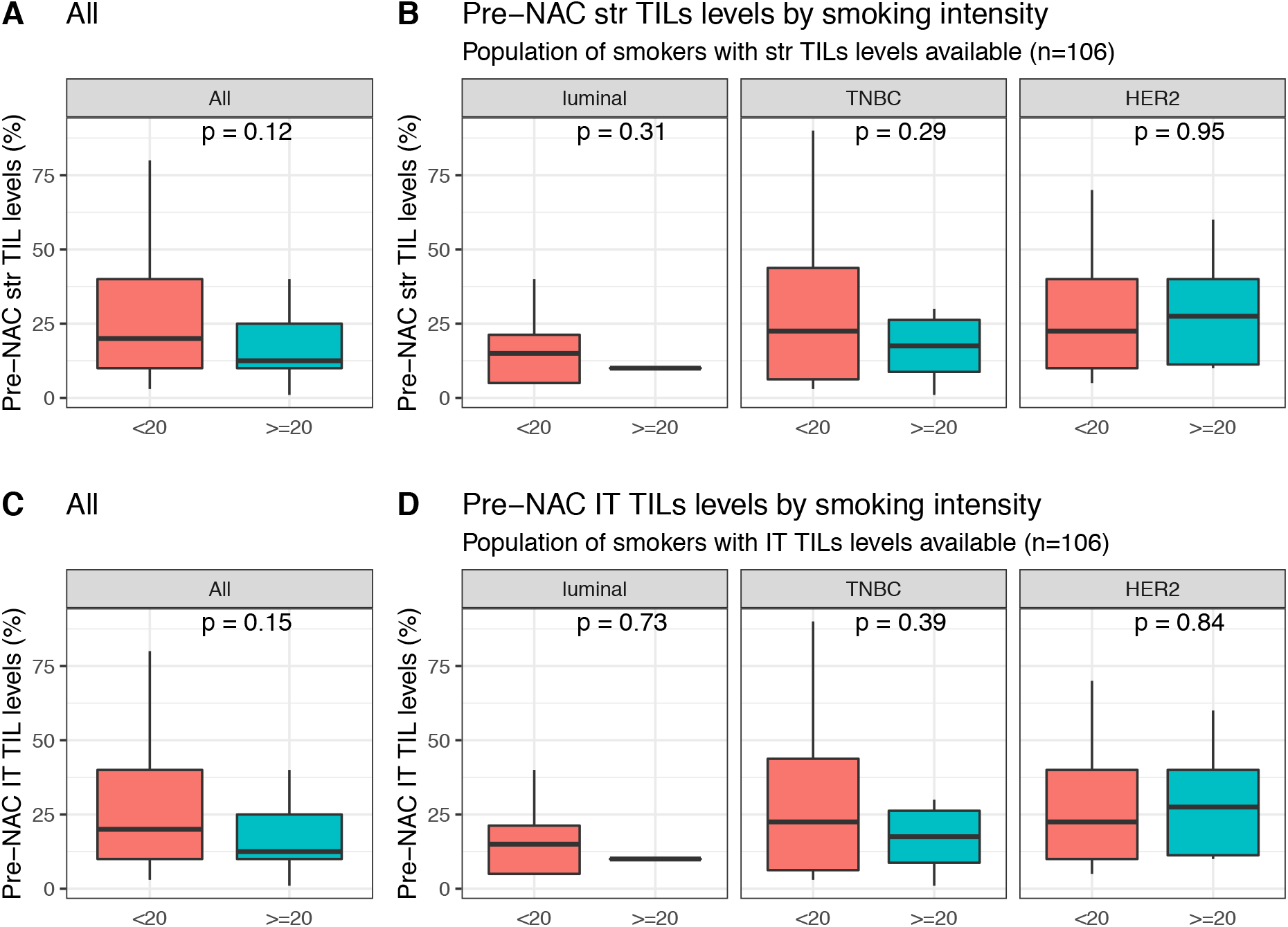
Pre-NAC TIL levels at BC diagnosis according to smoking amount (< 20 pack- years or ≥ 20 pack-years): Pre-NAC str TIL levels in the whole population (A), Pre-NAC str TIL levels by BC subtype (B), Pre-NAC IT TIL levels in the whole population(C), Pre-NAC IT TIL levels by BC subtype (D).

**Supplementary Figure 2:**
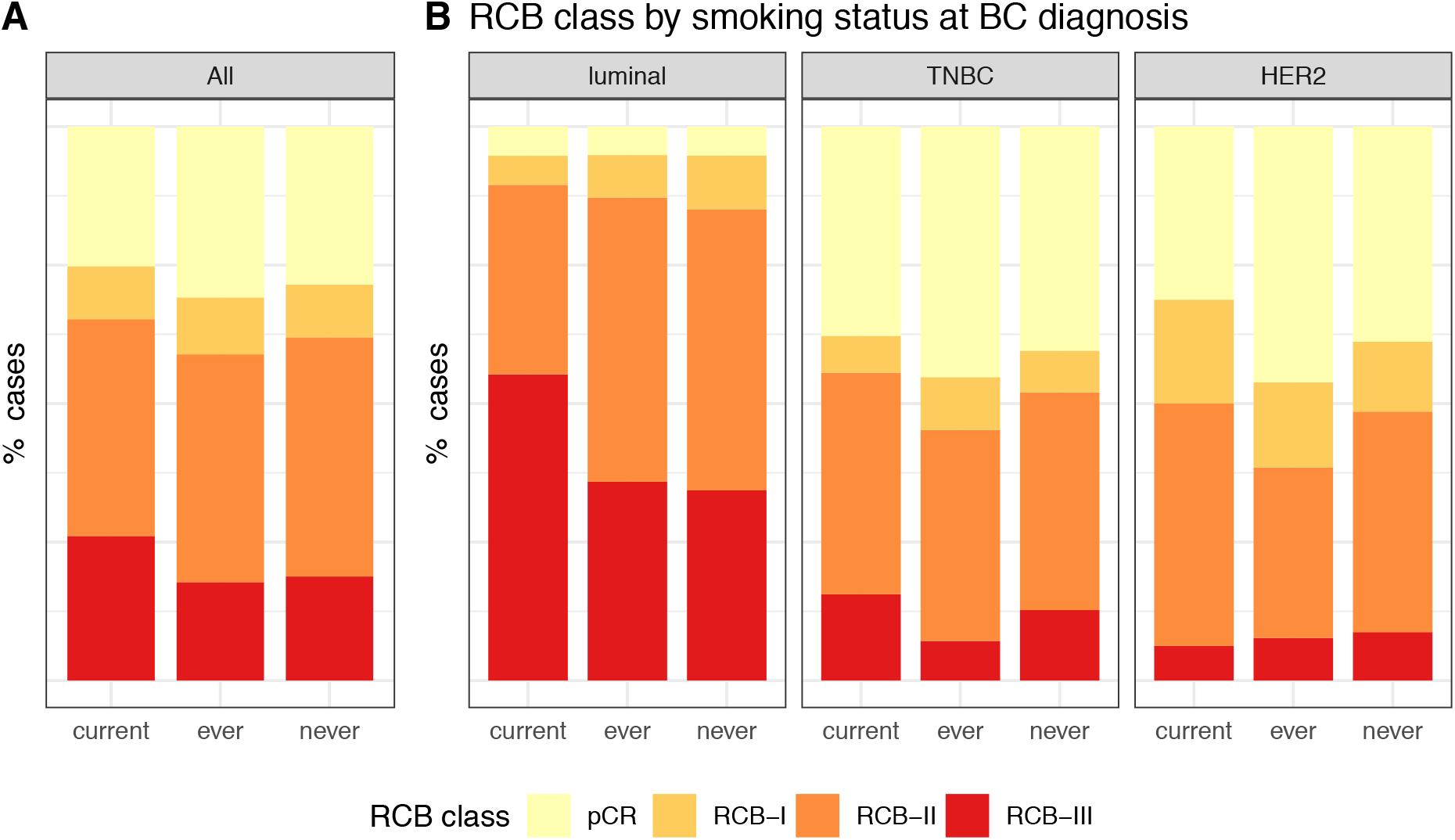
RCB class at NAC completion, according to smoking status, in the whole population (A) and by BC subtype (B).

## Notes

### Competing Interest Statement

The authors have declared no competing interest.

### Clinical Trial

cohort CNIL declaration number 1547270

### Funding Statement

We thank Roche France for financial support for the construction of the Institut Curie neoadjuvant database (NEOREP). Funding was also obtained from the Site de Recherche Integree en Cancerologie/Institut National du Cancer (Grant No. INCa-DGOS-4654). A-S Hamy-Petit was supported by an ITMO-INSERM-AVIESAN translational cancer research grant. The funders had no role in study design, data collection and analysis, decision to publish, or preparation of the manuscript.

### Author Declarations

The study was approved by the Breast Cancer Study Group of Institut Curie and was conducted according to institutional and ethical rules regarding research on tissue specimens and patients. Written informed consent from the patients was not required by French regulations.

